# Genetically proxied PCSK9 inhibition provides indication of lower prostate cancer risk: a Mendelian randomization study

**DOI:** 10.1101/2022.04.18.22274003

**Authors:** Si Fang, James Yarmolinsky, Dipender Gill, Caroline J Bull, Claire M Perks, the PRACTICAL consortium, George Davey Smith, Tom R Gaunt, Tom G Richardson

## Abstract

**Background:** Prostate cancer (PrCa) is the second most prevalent malignancy in men worldwide. Observational studies have linked the use of low-density lipoprotein cholesterol (LDL-c) lowering therapies with reduced risk of PrCa, which may potentially be attributable to confounding factors. In this study, we performed a drug target Mendelian randomization (MR) analysis to evaluate the association of genetically proxied inhibition of LDL-c lowering drug targets on risk of PrCa.

**Methods and Findings:** Single-nucleotide polymorphisms (SNPs) in and around *HMGCR, NPC1L1* and *PCSK9* genes associated with LDL-c (P<5×10^−8^) from the Global Lipids Genetics Consortium genome-wide association study (GWAS) (N=173,082) were used to proxy the therapeutic inhibition of these targets. Association estimates for the risk of total, advanced and early-onset PrCa were obtained from the PRACTICAL consortium. Replication was performed using genetic instruments from an LDL-c GWAS conducted on male UK Biobank participants of European ancestry (N=201,678), as well as instruments selected based on liver-derived gene expression and circulation plasma levels of targets. We also investigated whether putative mediators may play a role in findings for traits previously implicated in PrCa risk (i.e., lipoprotein a (Lp(a)), body mass index (BMI) and testosterone).

Applying MR using the inverse-variance weighted approach accounting for genetic correlations between instruments provided strong evidence supporting an effect of genetically proxied inhibition of PCSK9 (equivalent to a standard deviation (SD) reduction in LDL-c) on lower risk of total PrCa (odds ratio (OR)=0.84, 95% confidence interval (CI)=0.74 to 0.96, P=7.86×10^−3^) and early-onset PrCa OR=0.70, 95% CI=0.52 to 0.95, P=0.021. Analyses using male-stratified instruments provided consistent results. In contrast, there was little evidence of an association of genetically proxied HMGCR (OR=0.83, 95% CI=0.67 to 1.03, P=0.093) or NPC1L1 (OR=1.27, 95% CI=0.87 to 1.87, P=0.218) inhibition on PrCa risk.

Secondary analyses supported a genetically proxied effect of liver-specific *PCSK9* expression (OR=0.90, 95% CI=0.86 to 0.95, P=5.50×10^−5^) and circulating plasma levels of PCSK9 (OR=0.93 per SD reduction in PCSK9, 95% CI=0.87 to 0.997, P=0.04) on PrCa risk. Colocalization using eCAVIAR identified evidence (colocalization posterior probability=0.103) of a shared genetic variant (rs553741) between liver-derived *PCSK9* expression and PrCa risk. Moreover, genetically proxied PCSK9 inhibition was strongly associated with Lp(a) levels (Beta= -0.07, 95% CI= -0.10 to -0.03, P=1.44×10^−4^), but not BMI or testosterone, indicating a putative mediatory role of Lp(a).

**Conclusions:** Our study supports a strong association between genetically proxied inhibition of PCSK9 and a lower risk of total and early-onset PrCa. Further evidence from clinical studies is needed to confirm this finding as well as the putative mediatory role of Lp(a).

## Introduction

Prostate cancer is the second most commonly diagnosed malignancy in men globally with over 1.4 million new cases in 2020 [1]. Findings from the literature have provided conflicting evidence of a relationship between elevated low-density lipoprotein (LDL) cholesterol and prostate cancer risk. For instance, pre-clinical studies have suggested that high levels of extracellular LDL cholesterol (LDL-c) may promote the proliferation of prostate cancer cells [2, 3]. Conversely, observational studies have typically found there to be limited evidence of an association between LDL-c [4-7] and overall risk of prostate cancer, although there are some which have reported that LDL-c lowering medications may reduce the risk of prostate cancer incidence [5, 8-10]. Taken together these findings suggest that, although LDL-c may not directly contribute towards prostate tumorigenesis, biological pathways which regulate the biosynthesis and metabolism of LDL-c may influence prostate cancer risk through alternate mechanisms.

The use of human genetics to evaluate the efficacy and safety profiles of therapeutic targets is becoming increasingly popular in drug development, with recent evidence suggesting that targets with the support of genetics are approximately twice as likely to successfully make it to market [11]. Furthermore, the wealth of readily accessible data from genome-wide association studies (GWAS) means that these types of evaluations are typically inexpensive and quick to undertake. An approach to investigate genetic support for a target is Mendelian randomization (MR) [12-14], a causal inference technique which harnesses randomly segregated genetic variants as instrumental variables to proxy the perturbation of therapeutic targets [15, 16].

The application of MR to examine the genetically proxied effects of drug targets (referred to as ‘drug target MR’) has demonstrated the validity of this approach to corroborate findings from clinical trials as long as the underlying assumptions are met [17]. For example, the efficacy of lipid lowering drug-targets in reducing risk of coronary artery disease has been shown by previous MR studies, for therapies such as statins (which targets HMG-CoA reductase (*HMGCR*)), Proprotein convertase subtilisin/kexin type 9 (*PCSK9*) inhibitors and Ezetimibe (which targets Niemann-Pick C1-Like 1 (*NPC1L1*)) [18, 19]. Moreover, drug target MR analyses have provided evidence of adverse effects reported in trials, such as the effect of statins on increased risk of type 2 diabetes, as well as highlighting potential additional indications [20]. Recently, this approach has also been applied to suggest that statins may provide additional benefit towards the lowering of ovarian cancer risk [21].

In this study, we applied drug target MR to investigate the association between genetically proxied LDL-c lowering medications and risk of total, advanced and early-onset prostate cancer using data from the Prostate Cancer Association Group to Investigate Cancer Associated Alterations in the Genome (PRACTICAL) consortium [22]. Genetic instruments were oriented to proxy the effect of statins, PCSK9 inhibitors and Ezetimibe on lowering circulating LDL-c. The associations of genetically proxied LDL receptor (LDLR) mediated LDL-c and the genetically proxied levels of LDL-c on prostate cancer outcomes were also examined to assess whether evidence of the findings for inhibiting drug targets may be generalisable to the lowering of LDL-c, i.e., the on target effects of this class of drug. Moreover, analyses using genetic variants to investigate the levels of circulating proteins and liver-specific gene expression for targets were performed as sensitivity analyses and to triangulate evidence [23]. The effect of LDL-c lowering drugs, LDLR expression and circulating LDL-c on advanced prostate cancer and early-onset prostate cancer was also investigated. Finally, we evaluated the genetically proxied effects of LDL-c lowering drugs on several traits previously implicated to play a role in prostate cancer risk (body mass index (BMI) [24-26], lipoprotein A (Lp(a)) [27] and testosterone [28, 29]) to discern whether they may reside along the pathways between therapeutic targets and risk of prostate cancer.

## Methods

### Ethics statement

Our work involves the previously collected genetic sequencing and phenotype data of human participants in the UK Biobank cohort study. The North West Multi-centre Research Ethics Committee (MREC) gave ethical approval for the UK Biobank.

### Data sources

In our primary analysis, two-sample Mendelian randomization (MR) was applied to study the association of genetically proxied therapeutic inhibition for lipid-lowering drug targets PCSK9, NPC1L1 and HMGCR, as well as genetically proxied levels of LDLR and overall LDL-c, on the risk of prostate cancer.

Genetic instruments for drug targets were extracted from the summary statistics from the Global Lipids Genetics Consortium (GLGC) genome-wide association study (GWAS) on LDL-c levels [30] through the IEU Open GWAS project. Final instruments for the three lipid-lowering drug targets (and genetically proxied LDLR) were genetic variants robustly associated with LDL-c (based on P < 5×10^−8^ and a pairwise linkage disequilibrium (LD) r^2^ < 0.1 using a reference panel consisting of individuals of European ancestry from the 1000 Genomes Project Consortium [31]) and located within 100kb around *PCSK9* (Entrez Gene: 255738), *NPC1L1* (Entrez Gene: 29881), *HMGCR* (Entrez Gene: 3156) and *LDLR* (Entrez Gene: 3949) respectively. Additionally, genome-wide variants associated with LDL-c were used as instrumental variables for overall levels of LDL-c (based on P<5×10^−8^ and r^2^<0.001).

Given that genetic estimates on prostate cancer risk were derived in a male-only population, we repeated all analyses using male-stratified instruments identified by conducting a GWAS of LDL-c (ID: 30780) on 201,678 unrelated male participants of European ancestry in the UK Biobank study [32]. The levels of LDL-c were standardized to have a mean of 0 and standard deviation of 1 prior to analysis. BOLT-LMM was implemented to conduct the GWAS analysis with adjustment for age and genotyping chip [33]. Further details of the GWAS analysis pipeline has been described previously [34, 35]. After quality control and imputation, the GWAS results were clumped with two sets of different P-value and LD r^2^ thresholds as described above, i.e., (1) P < 5×10^−8^ and pairwise r^2^ < 0.1, or (2) P<5×10^−8^ and r^2^<0.001. Male-stratified genetic instruments for the five risk factors of interest were extracted from the clumped GWAS summary statistics using the same criterion described above.

For prostate cancer outcomes, summary statistics were obtained from GWAS meta-analyses on risks of overall prostate cancer (n=140,306 men including 79,194 cases and 61,112 controls), early-onset (≤55 years) prostate cancer (n=51,244 men including 6,988 cases and 44,256 controls) and advanced prostate cancer (n=73,475 men including 15,167 cases 58,308 controls) and from the Prostate Cancer Association Group to Investigate Cancer Associated Alterations in the Genome (PRACTICAL) consortium [22]. Advanced prostate cancer cases include individuals with either metastatic prostate cancer, a Gleason score of 8 or higher, prostate-specific antigen level greater than 100 ng/mL or prostate cancer related death [22]. All individuals involved in the prostate cancer GWAS analyses are of European ancestry.

In secondary analyses, *cis*-acting protein quantitative trait loci (*cis*-pQTL) for PCSK9 were used as the genetic instruments for circulating PCSK9 protein levels. PCSK9 *cis*-pQTLs were extracted from a publicly available GWAS on plasma levels of PCSK9 measured in 35,559 Icelanders [36] and clumped using a window of 100kbs around the *PCSK9* encoding region based on P<5×10^−8^ & r^2^<0.1 with the same reference panel as above. All LD clumping was performed using PLINK (v1.9). A similar protocol was applied to select instruments for *PCSK9* expression derived from liver tissue was applied using data (N=208) from the latest release (v8) of the Genotype-Tissue Expression (GTEx) project [37]. As no liver-tissue PCSK9 *cis*-expression quantitative trait loci (cis-eQTL) reached the P-value threshold for MR analyses (P<5×10^−8^), the top eQTL (rs553741) which provided the strongest statistical evidence (P=6.02×10^−8^) and F-statistic (F=26.9) was used to derive the Wald ratio estimate for MR analyses on prostate cancer outcomes.

### Statistical analysis

#### Two-sample Mendelian randomization

In the primary analysis, two-sample Mendelian randomization (MR) was applied to investigate the associations of genetically proxied inhibition of lipid lowering drug targets and overall LDL-c on the risk of prostate cancer. All analysis were performed using the *TwoSampleMR* R package (v0.5.6, https://github.com/mrcieu/TwoSampleMR). The instrument strength (F statistics) for LDL-c lowering drug targets and risk factors examined in this study were calculated using a formula previously described by Bowden *et al* [38].

In the primary drug target MR analysis, the association estimate for inhibition of PCSK9 and NPC1L1 were estimated using the random-effects inverse-variance weighted (IVW) model [39]. The MR estimate of HMGCR inhibition was estimated using the Wald ratio method as only one SNP (rs12916) was identified as the genetic instrument for this target from the GLGC GWAS. Considering the weak LD between genetic variants (r^2^<0.1) used as instrumental variables, IVW analyses were adjusted for LD matrices between instruments based on the same reference panel as above to ensure they were independent of one and other [40]. Iterative leave-one-out analyses were conducted for PCSK9 to identify the presence of any single variants which may be driving identified effects on the outcome. In replication analysis using UKB male-stratified genetic instruments, the effects from inhibiting PCSK9 and HMGCR were analysed using IVW accounting for genetic correlations whereas the effects from inhibition of NPC1L1 was estimated using Wald ratio based on rs2073547.

Next, we performed two-sample MR using the random-effects IVW method to investigate whether the association between genetically proxied LDL-c lowering drug targets and total prostate cancer risk may be attributed to the inhibition of LDLR or due to overall changes in LDL-c. When analysing the effects of LDL-c levels, the weighted median model (which allows up to half of the included SNPs to be pleiotropic and is less influenced by outliers) [41], the weighted mode model (which assumes that the most common effect is consistent with the true causal effect) [42], and the MR-Egger model (which provides an estimate of association magnitude allowing all SNPs to be pleiotropic) [43] were used as sensitivity analysis.

As a secondary analysis, two-sample MR were performed using PCSK9 *cis*-eQTL and *cis*-pQTL to further examine results identified in the primary analyses. We firstly estimated the association between genetically proxied plasma levels of PCSK9 and LDL-c using the random-effects IVW methods accounting for LD between genetic variants. A Wald ratio was calculated to estimate the association between genetically proxied *PCSK9* expression (instrumented using a single *cis*-eQTL) and prostate cancer outcomes. We applied the random-effects IVW method accounting for correlation structure between genetic variants to examine the associations between genetically proxied plasma PCSK9 (instrumented using *cis*-pQTLs) and prostate cancer outcomes. The pairwise LD correlation r^2^ between eQTL and pQTLs were calculated using LDmatrix from LDlink [44] based on the reference panel consisting of Utah Residents from North and West Europe (CEU) individuals.

#### Co-localization between eQTL and total prostate cancer risk

Given that single SNP MR analyses can be prone to high false discovery rates due to linkage disequilibrium between the instrument and proximal variants, we conducted co-localization analyses to identify evidence of shared causal variants between liver-derived *PCSK9* gene expression and risk of total prostate cancer. The eCAVIAR (v2.2) approach was applied to formally appraise evidence as a colocalization posterior probability (CLPP) [45]. The cut-off of CLPP to indicate evidence of a shared causal variant is >0.01 as proposed by the authors of this approach [45]. Analyses by eCAVIAR were based on genetic variants within 100kb around the lead cis-eQTL for PCSK9 in liver tissue (rs553741). We constructed a locus zoom plot using *gassocplot2* R package (https://github.com/jrs95/gassocplot2) to compare the eQTLs associated with liver-specific *PCSK9* expression (GTEx v8) and variants associated with risk of prostate cancer.

#### Contrasting the genetically proxied associations between lipid lowering drugs targets and risk factors of prostate cancer

To examine whether the associations between genetically proxied inhibition of drug targets and prostate cancer risk could be attributed to changes in BMI, Lp(a) or testosterone, we performed two-sample MR to investigate effects from drug targets on those risk factors as a sensitivity analysis using the same methods as above. The associations between genetic variants and BMI were extracted (1) from the Pulit *et al*. GWAS meta-analysis on BMI (n= 806,834) [46] when using GLGC variants, and (2) from the Locke *et al*. GWAS on BMI (n=339,224) [47] when analysed using UKB male-stratified genetic variants to avoid sample overlap. The associations between genetic instruments and Lp(a) were extracted from a GWAS on inverse rank normalised levels of Lp(a) in UKB participants (http://www.nealelab.is/uk-biobank/) from the IEU Open GWAS project. Moreover, the associations between genetic variants and testosterone levels were from male stratified GWAS on total and bioavailable testosterone levels (n= 199,569 and 184,205 respectively) in the UK Biobank. Random-effects IVW models were used with adjustment for the linkage disequilibrium between instruments. For drug targets which are instrumented using a single genetic variant, Wald ratio estimates were used to evaluate their associations with the outcome.

## Results

### Genetic variant selection

Characteristics of genetic variants used to proxy the inhibition of therapeutic targets PCSK9, NPC1L1 and HMGCR are presented in **Table 1**. Details of genetic variants used to proxy the levels of LDLR-mediated LDL-c and total LDL-c are in **S1 Table**. Male-stratified genetic instruments for the five risk factors are also presented in **S1 Table**. Details of the *cis*-acting protein quantitative trait loci (*cis*-pQTL) for plasma levels of PCSK9 protein and liver-derived *cis*-acting expression quantitative trait loci (*cis*-eQTL) data for *PCSK9* gene are listed in **S2 Table**. The F statistics of genetic instruments for all drug targets and risk factors assessed in this study, including the eQTL and pQTLs, ranged from 26.9 to 372.1, suggesting that the results are unlikely to be biased due to weak instruments [48].

**Table 1.** Characteristics of LDL-c associated genetic variants used to proxy the inhibition of PCSK9, NPC1L1, and HMGCR.

| Exposure | SNP | Effect allele | Other allele | Effect | 95% CI |  | P |
| --- | --- | --- | --- | --- | --- | --- | --- |
| PCSK9 | rs2479409 | A | G | -0.064 | -0.072 | -0.056 | 2.51E-50 |
|  | rs7552841 | C | T | -0.037 | -0.045 | -0.028 | 5.40E-15 |
|  | rs11591147 | T | G | -0.497 | -0.532 | -0.462 | 8.57E-143 |
|  | rs11583974 | G | A | -0.065 | -0.088 | -0.042 | 3.95E-09 |
|  | rs2479394 | A | G | -0.039 | -0.047 | -0.031 | 1.58E-19 |
|  | rs11206510 | C | T | -0.083 | -0.093 | -0.073 | 2.38E-53 |
|  | rs10888896 | G | C | -0.043 | -0.052 | -0.033 | 2.14E-14 |
|  | rs585131 | C | T | -0.064 | -0.074 | -0.054 | 2.70E-35 |
| NPC1L1 | rs2073547 | A | G | -0.049 | -0.058 | -0.039 | 1.92E-21 |
|  | rs1434961 | C | T | -0.026 | -0.034 | -0.019 | 8.39E-12 |
| HMGCR | rs12916 | T | C | -0.073 | -0.081 | -0.066 | 7.79E-78 |

Genetic variants for each drug target were selected as weakly correlated (linkage disequilibrium $r^2 < 0.1$ ), genome-wide significant ( $P < 5 \times 10^{-8}$ ) single nucleotide polymorphisms (SNPs) associated with the levels of LDL-c and are located within 100kb around the respective encoding gene region. 95% CI: 95% confidence interval. Effects are in standard deviation change of LDL-c levels per copy of the effect allele.

### Mendelian randomization analysis of lipid lowering therapies and prostate cancer risk

We firstly applied drug target MR to investigate the associations of genetically proxied lipid-lowering drug targets (HMGCR, PCSK9 & NPC1L1) with overall prostate cancer risk (**Figure 1A**; **S3 Table**). Genetically proxied inhibition of PCSK9 was strongly associated with a lower risk of developing prostate cancer (IVW MR odds ratio (OR)=0.84, 95% confidence interval (95% CI) = 0.74 to 0.96, P=0.008, per standard deviation (SD) reduction in LDL-c). Leave-one-out analyses provided consistent evidence of an association between genetically proxied PCSK9 inhibition and risk of prostate cancer, suggesting that the overall estimate was not driven by a single influential variant (**Figure 1B**; **S4 Table**). Genetically proxied inhibition of HMGCR provided evidence of a similar magnitude of association with overall prostate cancer as PCSK9, although the 95% confidence interval included the null (OR=0.83, 95% CI=0.67 to 1.03, P=0.093). There was little evidence of an association between genetically proxied inhibition of NPC1L1 with overall prostate cancer risk (Wald ratio: OR=1.27, 95% CI=0.87 to 1.87, P=0.218). MR analysis on the risk of early-onset and advanced prostate cancer identified strong evidence for an association between genetically proxied inhibition of PCSK9 and early-onset disease (OR=0.70, 95% CI=0.52 to 0.95, P=0.021), but not with advanced prostate cancer (OR=0.96, 95% CI=0.79 to 1.17, P=0.703) (**S3 Table**). Similar findings were observed using male-stratified genetic instruments identified using the UKB data (**S3 Table**).

**Figure 1.**
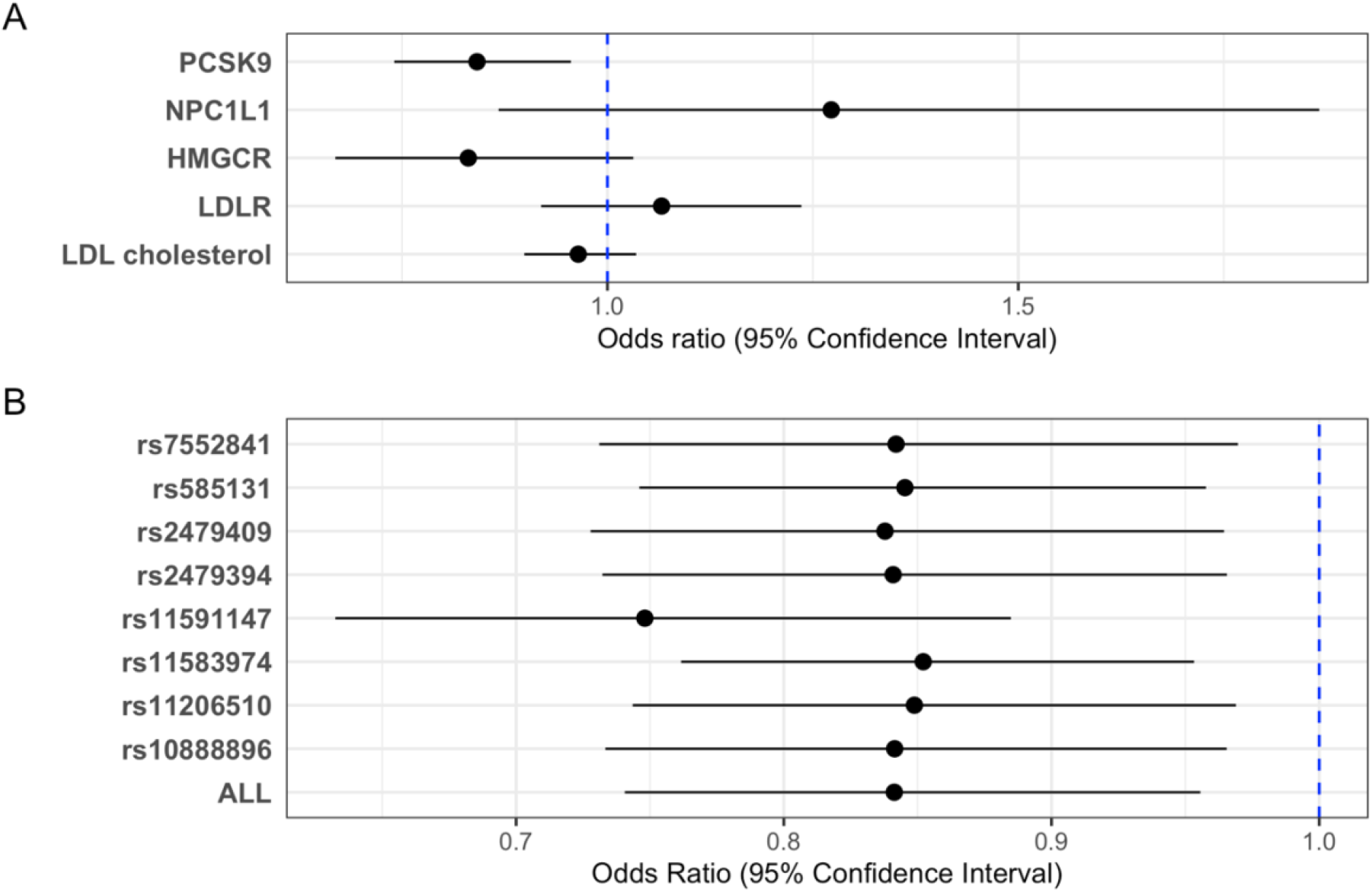
Results from Mendelian randomization analyses to estimate the effect of lipid lowering therapies and risk factors on overall prostate cancer risk. (A) Estimates of effects from genetically proxied inhibition of PCSK9, NPC1L1, HMGCR, LDLR and overall LDL-c on the overall risk of prostate cancer. (B) Leave-one-out Mendelian randomization results for the association between genetically proxied inhibition of PCSK9 and overall prostate cancer risk.

For comparative purposes, we investigated the effect of circulating LDL-c on overall prostate cancer risk by selecting genetic instruments at the *LDLR* locus as well as those associated with LDL-c across the genome. There was minimal evidence to suggest an effect of genetically proxied LDLR mediated LDL-c levels on overall prostate cancer risk (OR=1.07 equivalent to a SD reduction in LDL-c, 95% CI=0.92 to 1.124 P=0.398). Similarly, a 77 SNP genetic instrument for circulating LDL-c was minimally associated with prostate cancer risk (OR=0.96 equivalent to a SD reduction in LDL-c, 95% CI=0.90 to 1.04 P=0.316). Estimates for (overall) prostate cancer risk were replicated using a male-specific 103 SNP LDL-c instrument based on UK Biobank summary statistics, (OR=0.95 equivalent to a SD reduction in LDL-c, 95% CI=0.89 to 1.01, P=0.093). MR-Egger, weighted median and weighted mode estimates were comparable to the IVW results (**S3 Table**).

### Triangulation of evidence using data on circulating protein levels and liver-derived gene expression

To further examine the association between PCSK9 and prostate cancer, two-sample MR analyses were performed using *cis*-acting pQTLs to instrument inhibition of circulating PCSK9 protein levels (**S5 Table**). These *cis*-pQTLs are strongly associated with LDL-c levels (Beta=-0.54 SD change in LDL-c per SD reduction in PCSK9 levels, 95% CI=-0.59 to -0.49, P=3.81×10^−90^). MR results provided evidence of an effect of PCSK9 inhibition on overall prostate cancer (OR=0.93 per SD reduction in PCSK9 levels, 95% CI=0.87 to 0.997, P=0.040), and early-onset prostate cancer (OR=0.86, 95% CI=0.74 to 0.98, P=0.030) but not advanced prostate cancer (OR=0.98, 95% CI=0.89 to 1.07, P=0.600) using the IVW method accounting for genetic correlation structure, consistent with findings from our initial analyses.

LDL-c removal occurs primarily in the liver, which is also the organ where *PCSK9* is predominantly expressed based on the latest release (v8) of the Genotype-Tissue Expression (GTEx) project [37]. Based on our variant selection criteria (near PCSK9 gene and pairwise r^2^<0.1) and a false-discovery rate threshold of 0.05 defined by GTEx, there was only one eQTL in liver tissue using this dataset (rs553741), which is in strong LD with one of the SNPs used as genetic instruments (rs472495) to proxy the effects of PCSK9 inhibition in the primary drug target MR (r^2^ = 0.909) (for full LD matrix see **S6 Table**). MR estimates were supportive of an association between lower levels of *PCSK9* gene expression (instrumented using the eQTL) and a lower risk of overall prostate cancer (OR=0.90 per 1-SD reduction in *PCSK9* transcript levels, 95% CI=0.86 to 0.95, P=5.50×10^−5^). Analysis on disease subtypes provided similar magnitude of association with genetically proxied PCSK9, although confidence intervals overlapped the null (early onset prostate cancer: OR=0.91, 95% CI=0.91 to 1.03, P=0.139; advanced prostate cancer: OR=0.93, 95% CI=0.86 to 1.01, P=0.102).

As MR analyses using single variant instruments are particularly susceptible to false positive findings due to linkage disequilibrium structure with nearby genes [49], we conducted genetic colocalization at this locus to evaluate evidence of a shared causal variant between *PCSK9* expression in liver tissue and prostate cancer. Using the eCAVIAR method [45], we found a colocalization posterior probability (CLPP) of 0.103 for the variant rs553741, which suggests there is strong evidence of a shared causal variant at this locus based on a threshold of CLPP>0.01 as proposed by the authors of this approach [45]. Detailed CLPP for every candidate SNP are presented in **S7 Table**. A locus zoom plot comparing the *cis*-acting eQTLs associated with liver specific *PCSK9* expression and SNPs associated with risk of prostate cancer is shown in **Figure 2**. This evidence suggests that *PCSK9* gene expression in the liver and prostate cancer risk share a common causal variant.

**Figure 2.**
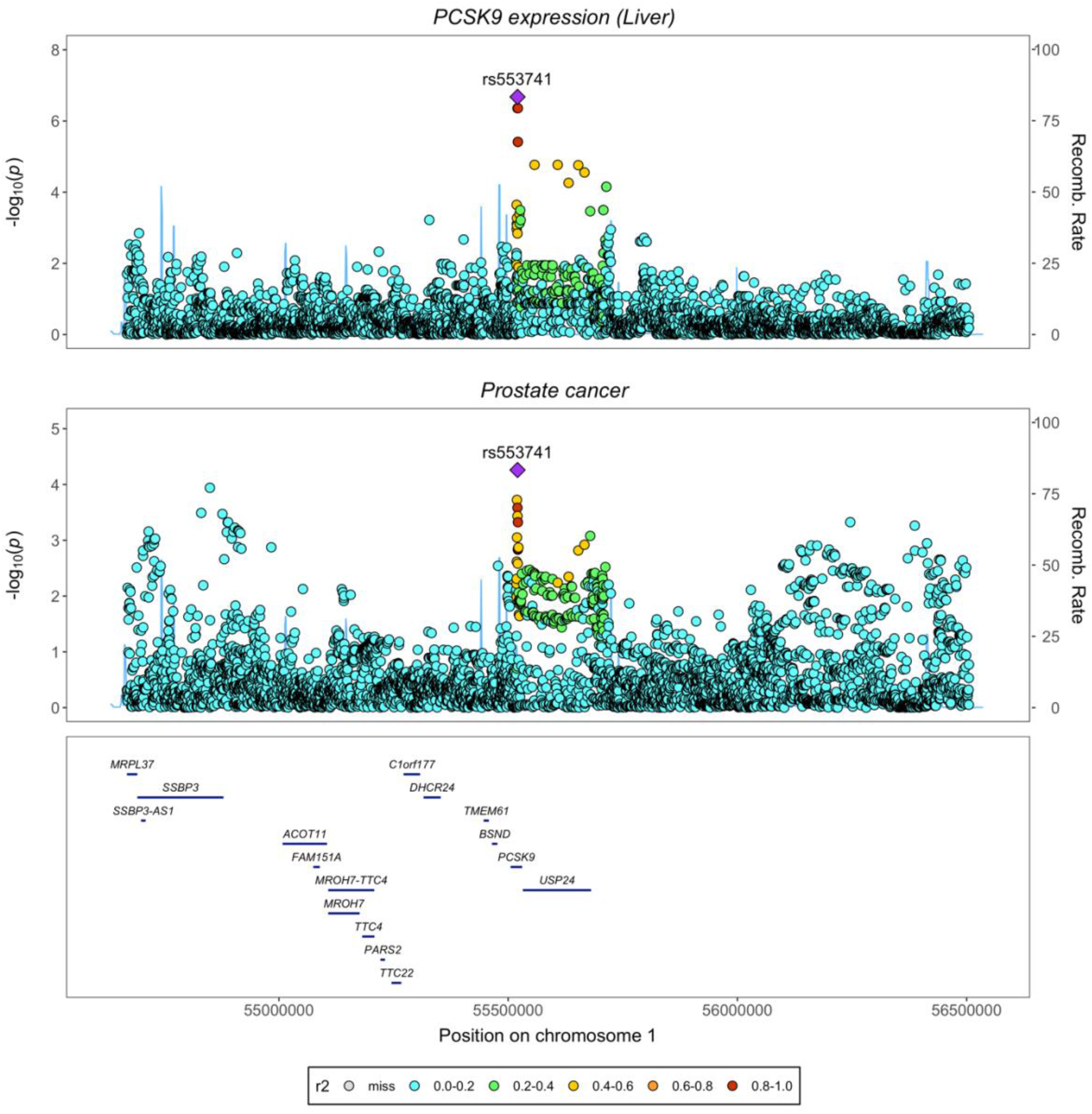
Locus zoom plots illustrating evidence of genetic colocalization between *PCSK9* gene expression in the liver and prostate cancer risk at the *PCSK9* gene locus.

### Contrasting the genetically proxied associations between lipid lowering drug targets and risk factors of prostate cancer

The association between genetically proxied lipid lowering drug target inhibition and prostate cancer may be mediated through prostate cancer risk factors, such as BMI, Lp(a) or testosterone. Therefore, we examined the association between genetically proxied inhibition of drug targets and risk factors for prostate cancer using drug target MR (**Figure 3**).

**Figure 3.**
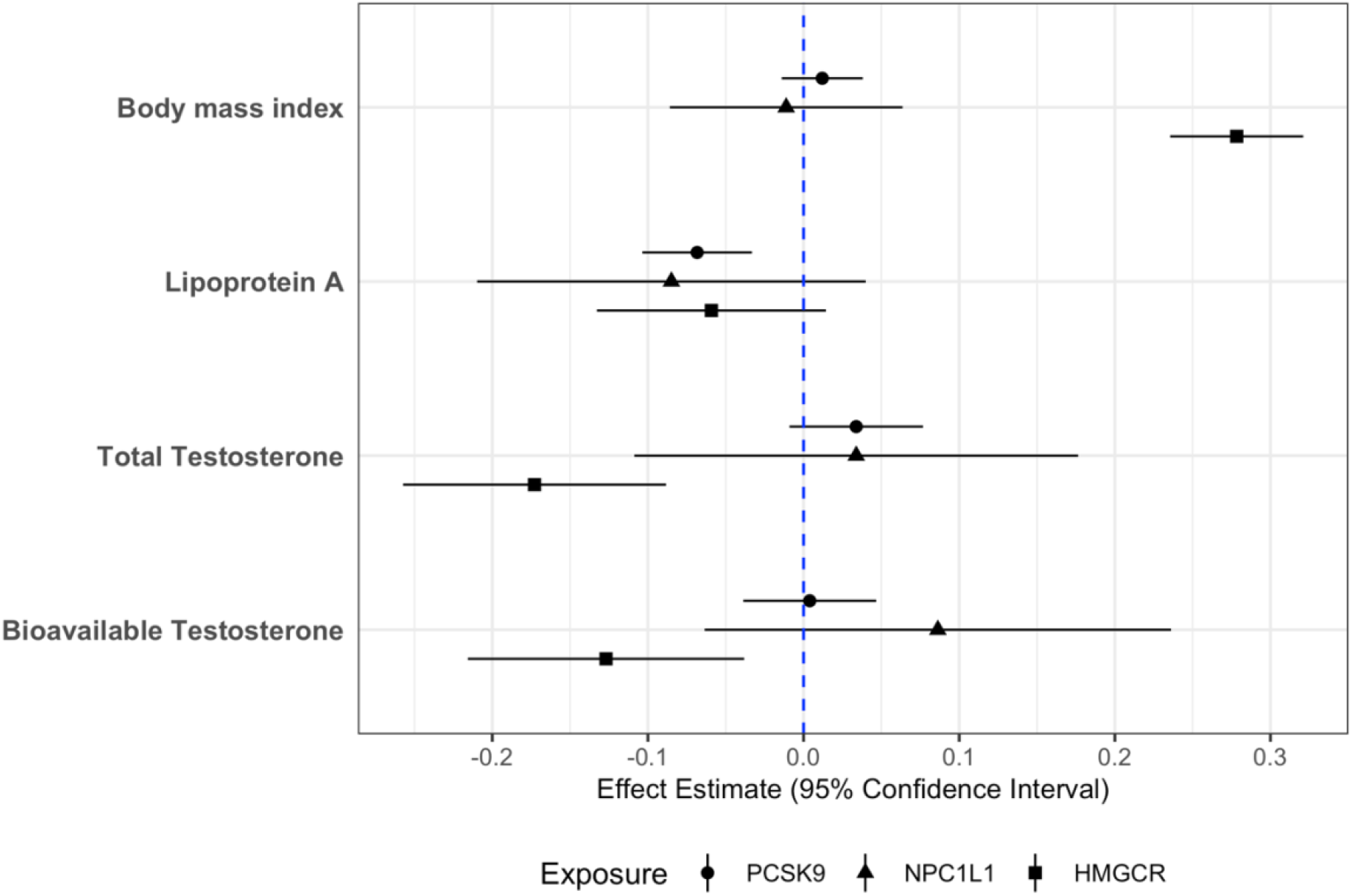
Results from drug target Mendelian randomization (MR) analyses to investigate the effect of lipid lowering therapies on body mass index, lipoprotein A and testosterone. Estimates of effects from genetically proxied inhibition of PCSK9, NPC1L1 and HMGCR on body mass index, levels of lipoprotein A, total testosterone, and bioavailable testosterone. Effect estimates are in standard deviation change in the outcome per drug target inhibition effect equivalent to a standard deviation reduction in LDL-c. Results were from analysis using instruments identified from the Global Lipids Genetics Consortium data.

Repeating our primary MR analyses to investigate the genetically proxied association of each lipid lowering target on BMI (**S8 Table**) provided little evidence for inhibition of PCSK9 (Beta=0.01 SD change in BMI per SD reduction in LDL-c, 95% CI=-0.01 to 0.04, P= 0.336) as well as NPC1L1 (Wald ratio: Beta=-0.01, 95% CI=-0.09 to 0.06, P= 0.770). However, genetically proxied inhibition of HMGCR provided strong evidence for an association with elevated body mass index (Wald ratio: Beta=0.28, 95% CI=0.24 to 0.32, P=3.12×10^−37^).

Evaluating effects on Lp(a) for each target (**S9 Table**) suggested that there was strong evidence of an effect of PCSK9 inhibition on lower levels of this lipoprotein particle (IVW accounting for LD matrix: Beta=-0.10 SD change in the levels of Lp(a) per 33.4 mg/dl reduction in LDL-c, 95% CI=-0.15 to -0.05, P=4.90×10^−5^). The association was supported in an analysis using PCSK9 pQTLs as the genetic instruments (Beta=-0.03 SD change in the levels of Lp(a) per SD reduction in plasma PCSK9 levels, 95%CI=-0.05 to -0.02, P=1.47×10^−4^). Conversely, investigating the effects of genetically proxied inhibition of HMGCR (IVW accounting for LD matrix: Beta=-0.09, 95% CI=-0.19 to 4.91×10^−3^, P=0.063) and NPC1L1 (Wald ratio: Beta=-0.21, 95% CI=-0.44 to 0.02, P=0.080) on Lp(a) levels found that their confidence intervals included the null despite similar central magnitudes of effect compared with the PCSK9 score. The putative causal relationship identified in this analysis is illustrated in a directed acyclic graph (**Figure 4**).

**Figure 4.**
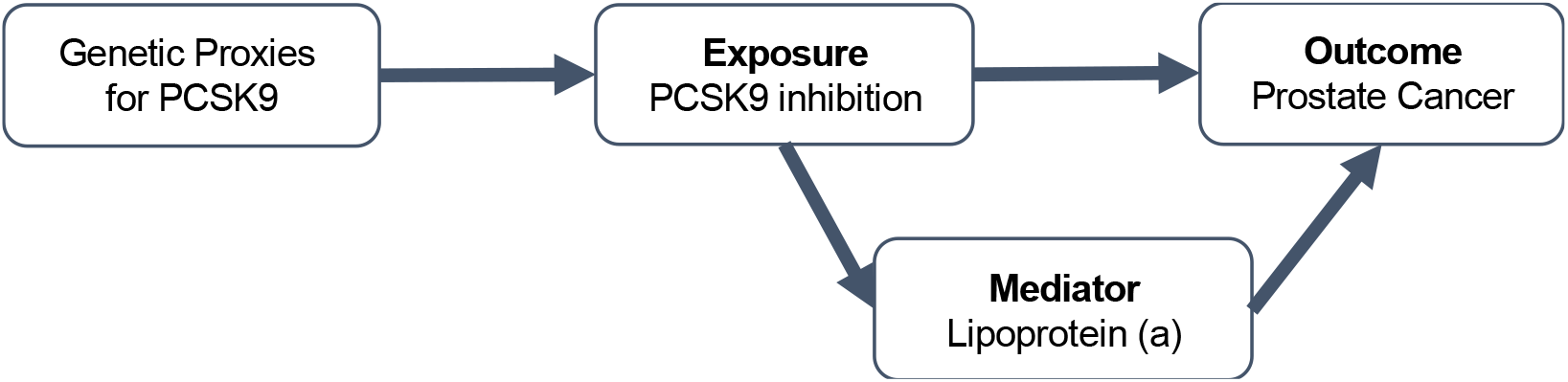
A directed acyclic graph showing the putative association between PCSK9 inhibition, lipoprotein A and prostate cancer.

Examining the effects from drug targets on testosterone levels (**S10 Table**) suggest genetically proxied inhibition of neither PCSK9 or NPC1L1 contributed to alterations in total testosterone (PCSK9: Beta=0.03 SD change in testosterone per drug effect equivalent to a SD reduction in LDL-c, 95% CI=-0.01 to 0.08, P=0.122; NPC1L1: Beta=0.03, 95% CI=-0.11 to 0.18, P=0.642) or bioavailable testosterone (PCSK9: Beta=3.97×10^−3^, 95% CI=-0.04 to 0.05, P=0.855; NPC1L1: Beta=0.09, 95%CI=-0.06 to 0.24, P=0.259) in men. On the contrary, genetic variants proxying the inhibition of HMGCR showed strong correlation with both measurements of testosterone (total: Beta=-0.26, 95% CI=-0.37 to -0.15, P=3.34×10^−6^; bioavailable: Beta=-0.13, 95% CI=-0.22 to -0.04, P=5.08×10^−3^) in men.

## Discussion

In this work, we have identified novel evidence using large-scale genetic data to suggest that therapeutic inhibition of lipid lowering drug target PCSK9 may reduce prostate cancer risk. Estimates based on circulating PCSK9 protein data and *PCSK9* expression data derived from liver tissue, where PCSK9 is primarily synthesized and secreted in the body, further support this finding. In contrast, LDL-c associated variants located at the *NPC1L1, HMGCR* and *LDLR* genetic loci, as well as the genetically proxied levels of circulating LDL-c, were not associated with the risk of prostate cancer. Taken together, these findings suggest that the genetically proxied association between PCSK9 inhibition and a lower risk of prostate cancer is unlikely to be due to a mechanism involving the lowering of LDL-c levels. We postulate that one potential explanation for this finding is due to the lowering of Lp(a), which genetically proxied PCSK9 inhibition provided stronger evidence of achieving in comparison to statin and Ezetimibe therapies in this study.

PCSK9 is known as a regulator of the metabolism of LDL-c, although thus far its role in cancer susceptibility has yet to be comprehensively evaluated and characterized. There are various pre-clinical studies based on tumour tissue or mouse models which report the direct effects of PCSK9 or PCSK9 inhibitors on multiple types of cancer [50], including hepatocellular carcinoma [51, 52], lung carcinoma [53, 54], colorectal cancer [55] and breast cancer [56]. Additionally, Liu *et al*. inoculated *Pcsk9* knockout mouse cancer cells into syngeneic mouse hosts and observed delayed tumour growth, as well as a synergistic effect between PCSK9 inhibition and anti-PD1 antibody treatment to promote the efficacy of tumour growth suppression [57]. In contrast, few pre-clinical studies have linked PCSK9 to prostate cancer [58, 59], although Gan *et al*. previously demonstrated that *PCSK9* siRNA protects human prostate cancer cells from ionizing radiation induced cell damage [60].

With excellent efficacy and safety profiles, two monoclonal antibodies against PCSK9 have been developed to lower elevated LDL-c levels and subsequently help prevent coronary heart disease (CHD) [61, 62]. Our findings add to growing evidence suggesting that PCSK9 inhibitors may provide the most benefit towards reducing disease risk in comparison to statins and Ezetimibe which inhibit HMGCR and NPC1L1, respectively.

Furthermore, recent evaluations involving CRISPR base editing in primates suggests that complete knockdown of PCSK9 in the liver results in approximately a 60% reduction of LDL cholesterol [63]. Further research is required to investigate the consequences of this approach towards prostate cancer risk, although our findings using genetic proxies of PCSK9 inhibition predict that this would have a beneficial effect, especially on early-onset prostate cancer. This supports the recently identified favourable overall effects from genetically proxied PCSK9 inhibition on the lifespan [64]. In addition, prostate cancer, like various other types of cancer, undergoes multiple stages of accumulations of oncogenic mutations to develop into advanced and eventually metastatic disease, which is highly lethal and heterogenous [65]. The lack of evidence for an association between PCSK9 inhibition and the risk of advanced prostate cancer suggest circulating PCSK9 is less likely to be promoting the progression of the disease.

We conducted various sensitivity analyses to further investigate the finding between genetically proxied PCSK9 inhibition and prostate cancer risk. For instance, leave-one-out analysis found consistent effect estimates for genetically proxied PCSK9 on prostate cancer risk, suggesting the association is unlikely to be driven by any single variant in our instrument. This mitigates the likelihood that individual pleiotropic variants at the *PCSK9* locus are influencing prostate cancer risk via alternate biological pathways. Moreover, we have triangulated evidence from plasma protein and liver-derived gene expression data to further support findings from our primary MR approach. Further evidence from genetic colocalization analyses using liver-derived gene expression data suggests that our finding is unlikely to be explained by a pathway involving a neighbouring gene as opposed to *PCSK9*.

Additionally, we performed subsequent analyses to explore the potential mediatory role of BMI, Lp(a) and testosterone in the associations between drug targets and prostate cancer risk. Elevated BMI has been found to associated with increased prostate cancer risk in multiple observational studies [24-26], whereas MR study identified evidence for an inversed association between them [66]. In addition, levels of testosterone were found to be strongly correlated with the risk of prostate cancer in both observational [28] and MR [29] studies. Our results found little evidence for associations for mediatory effects from genetically proxied inhibition of PCSK9 on these two risk factors. Moreover, a higher levels of Lp(a) has previously been reported to associate with poor prognosis of prostate cancer [67] as well as a higher risk of overall and early onset prostate cancer [27]. Our results provided strong evidence that PCSK9 inhibition may lower levels of Lp(a), and there is weak evidence for a similar effect from the inhibition of NPC1L1 or HMGCR. This finding has also been supported by several randomized control trials [61, 68] which found that PCSK9 inhibitors significantly reduced levels of Lp(a), whereas statins and Ezetimibe had little to mild effects on Lp(a) [69]. Furthermore, genetically proxied PCSK9 exhibited the strongest magnitude of association with the risk of early onset prostate cancer than that of overall prostate cancer, and there is weak evidence for association with advanced disease. This is consistent with the association between Lp(a) and the risk of prostate cancer outcomes identified in the recent multivariable MR [27]. Although our findings suggest that Lp(a) may play a mediatory role along the pathway between PCSK9 inhibition and prostate cancer risk, further functional work is required to robustly demonstrate this.

This study has noteworthy limitations. Firstly, although an SD shift in LDL-c is clinically achievable, the scale of the estimate identified for genetic proxies of therapeutic targets may not be equivalent to those reported by randomized controlled trials: whilst drugs are often taken for a defined period, MR analyses are conventionally believed to provide an estimate considering lifelong exposure to risk factors, with increasing evidence suggest the associations between genetic instruments and the exposure may vary throughout the life course [70]. Additionally, conventional MR methods assume a linear relationship between genetically proxied exposures and outcomes, however, drugs may not trigger any biological response until a drug dose exceeds a certain level. Secondly, we leveraged 13 *cis*-pQTLs to instrument plasma levels of PCSK9 in this work due to the availability of a large-scale dataset for whole blood measures (n=35,559) [36]. However, analyses using *cis*-pQTL derived from liver tissue would be valuable to further examine the effect of PCSK9 inhibition on prostate cancer risk once these data are available in sufficient samples. Thirdly, using genetic instruments at PCSK9 locus extracted from a GWAS of LDL-c, liver derived PCSK9 expression and circulating PCSK9 protein, this work focuses on the indirect association between PCSK9 inhibition and prostate cancer risk. Future studies with prostate derived PCSK9 *cis*-pQTL are essential for evaluating the direct role of PCSK9 in prostate cancer cells, especially the effect on advanced prostate cancer. Furthermore, replication of MR estimates using males-stratified PCSK9 *cis*-eQTL and *cis*-pQTL would be worthwhile to support this finding in the future, even though we did not find large differences using LDL-c stratified instruments to overall conclusions.

In summary, our study demonstrates that genetically proxied inhibition of PCSK9 is strongly associated with a lower risk of overall and early onset prostate cancer, potentially through a mechanism involving the lowering of Lp(a) levels. Further evidence from clinical studies on prostate cancer incidence and progression among patients taking PCSK9 inhibitors is needed to confirm this finding.

## Data Availability

Several summary statistics of genome wide association studies (GWAS) used in this study are available on the IEU Open GWAS project (https://gwas.mrcieu.ac.uk/), including LDL cholesterol GWAS from the Global Lipids Genetics (GLGC) Consortium, overall prostate cancer risk GWAS from the Prostate Cancer Association Group to Investigate Cancer Associated Alterations in the Genome (PRACTICAL) consortium, Locke et al. GWAS on body mass index, as well as GWAS on lipoprotein A, male-stratified total and bioavailable testosterone from the Neale lab. Pulit et al. GWAS summary statistics on body mass index is available from https://zenodo.org/record/1251813#.YlhVzZPMJhE. Liver-derived PCSK9 eQTL data is available from the GTEx project via https://www.gtexportal.org/home/. PCSK9 plasma pQTL data is available from https://www.decode.com/summarydata/. Summary statistics of GWAS on advanced and early onset prostate cancer risks are under restricted access. They are available from the PRACTICAL consortium upon application (contact). The summary statistics of GWAS on male-stratified LDL cholesterol in the UK Biobank will be available after acceptance.

## Acknowledgement

The Genotype-Tissue Expression (GTEx) Project was supported by the Common Fund of the Office of the Director of the National Institutes of Health, and by NCI, NHGRI, NHLBI, NIDA, NIMH, and NINDS. The data used for the analyses described in this manuscript were obtained from: the GTEx Portal on 12/12/2021. See the separate file for an extended list of acknowledgements for the PRACTICAL consortium, CRUK, BPC3, CAPS, PEGASUS.

## Supporting information captions

**S1 Table. Genetic variants used as instrumental variables for drug targets and LDL-c levels in Mendelian randomization analysis**.

(See separate excel file)

**S2 Table. Genetic variants (liver pQTLs) used as instrumental variables for plasma levels of PCSK9 protein**.

(See separate excel file)

**S3 Table. Results from Mendelian randomization investigating the associations between genetically proxied inhibition of drug targets, reduction of LDLR levels of LDL-c levels and the risk of prostate cancer**. Estimates are effects equivalent to 1 standard deviation reduction in LDL-c levels.

(See separate excel file)

**S4 Table. Results from sensitivity tests on the PCSK9 drug target Mendelian randomization on total prostate cancer risk**. These analyses were performed using genetic instruments identified from the Global Lipids Genetics Consortium (GLGC).

(See separate excel file)

**S5 Table. Results from Mendelian randomization investigating the associations between plasma PCSK9 pQTL and liver derived PCSK9 eQTL and prostate cancer outcomes**. Estimates are per 1 standard deviation reduction in plasma protein levels of PCSK9 for pQTLs or per 1 standard deviation reduction in the levels of PCSK9 transcripts in the liver tissue.

(See separate excel file)

**S6 Table. Pairwise linkage disequilibrium correlation coefficients between liver derived PCSK9 *cis*-eQTL and plasma PCSK9 pQTLs**. The pairwise r^2^ were calculated using the reference panel consisting of Utah Residents from North and West Europe (CEU) individuals.

(See separate excel file)

**S7 Table. Colocalization posterier probability (CLPP) of candidate causal genetic variants shared between PCSK9 gene expression in liver and prostate cancer risk**.

(See separate excel file)

**S8 Table. Results from Mendelian randomization analysis on the associations between genetically proxied inhibition of drug targets, reduction of LDLR levels of LDL-c levels and body mass index**. Estimates are effects equivalent to 1 standard deviation reduction in LDL-c levels or 1 standard deviation reduction in plasma protein levels of PCSK9.

(See separate excel file)

**S9 Table. Results from Mendelian randomization analysis on the associations between genetically proxied inhibition of drug targets, reduction of LDLR levels of LDL-c levels and the levels of lipoprotein A**. Estimates are effects equivalent to 1 standard deviation reduction in LDL-c levels or 1 standard deviation reduction in plasma protein levels of PCSK9.

(See separate excel file)

**S10 Table. Results from Mendelian randomization analysis on the associations between genetically proxied inhibition of drug targets, reduction of LDLR levels of LDL-c levels and the levels of testosterone**. Estimates are effects equivalent to 1 standard deviation reduction in LDL-c levels or 1 standard deviation reduction in plasma protein levels of PCSK9.

(See separate excel file)

